# Sex steroid hormones and risk of breast cancer incidence and survival: A two-sample Mendelian randomization study

**DOI:** 10.1101/2021.10.14.21264952

**Authors:** Aayah Nounu, Siddhartha P Kar, Caroline L Relton, Rebecca C Richmond

## Abstract

**Background:** Breast cancer (BC) is the cancer with the highest incidence and mortality in women worldwide. Observational epidemiological studies suggest a positive association between testosterone, estradiol, dehydroepiandrosterone sulphate (DHEAS) and both pre- and post-menopausal BC. Since previous studies may be prone to bias and confounding, we used a two-sample Mendelian randomization (MR) analysis to investigate this association.

**Methods:** Genetic instruments for nine sex steroid hormones and sex hormone binding globulin (SHBG) were obtained from genome-wide association studies (GWAS) conducted in the UK Biobank (total testosterone (TT) N:230,454, bioavailable testosterone (BT) N: 188,507 and SHBG N: 189,473), The United Kingdom Household Longitudinal Study (DHEAS N: 9,722), the LIFE-Adult and LIFE-Heart cohorts (estradiol N: 2,607), the LIFE-Heart cohort only (androstenedione N: 711, aldosterone N: 685, progesterone N: 1,259 and 17-hydroxyprogesterone N: 711) and the CORtisol NETwork (CORNET) consortium (cortisol N: 25,314). GWAS summary statistics were also obtained from the Breast Cancer Association Consortium (BCAC) for overall BC risk (N: 122,977 cases and 105,974 controls) and BC mortality (96,661 BC cases and 7,697 BC-specific deaths). Subtype specific analysis were carried out for incidence of estrogen receptor (ER)+ BC, ER- BC, luminal A-like BC, luminal B-like BC, luminal B/HER2-negative-like BC, HER2-enriched-like BC, triple negative BC (TNBC) and *BRCA1* mutated TNBC.

**Results:** Using an inverse-variance weighted (IVW) approach, we found that a standard deviation (SD) increase in TT, BT and estradiol increased the risk of overall BC (OR: 1.14, 95% CI: 1.09-1.21, OR: 1.19, 95% CI: 1.07-1.33 and OR: 1.03, 95% CI: 1.01-1.06, respectively) and ER+ BC (OR: 1.19, 95% CI: 1.12-1.27, OR: 1.25, 95% CI: 1.11-1.40 and OR: 1.06, 95% CI: 1.03-1.09, respectively). A SD increase in DHEAS also increased ER+ BC risk (OR: 1.09, 95% CI: 1.03-1.09). Subtype specific analyses showed similar associations with ER+ expressing subtypes: luminal A-like BC, luminal B-like BC and luminal B/HER2-negative-like BC. A SD increase in cortisol was associated with poor survival after a diagnosis of ER-BC (HR: 2.35, 95% CI: 1.00-5.49).

**Discussion/Conclusion:** TT, BT, DHEAS and estradiol increase the risk of ER+ type BCs similar to observational studies, but none of these hormone measures are associated with BC survival. We found some evidence that cortisol reduced ER- BC survival. Stronger genetic instruments are required before definitive conclusions can be made about the role of other sex-steroid hormones in breast cancer. Understanding the role of sex steroid hormones in BC risk, particularly subtype-specific risks, highlights the potential importance of attempts to modify and/or monitor hormone levels in order to prevent BC.

## Introduction

Breast cancer (BC) is the most common cancer in women worldwide and is the leading cause of cancer mortality in females (1). Early menarche and a later age at menopause have been shown to be associated with an increased risk of breast cancer (2). Furthermore, a study conducted in postmenopausal women showed that a higher number of lifetime cumulative menstrual cycles increased BC risk (3). Taken together, susceptibility to BC appears to be associated with ovarian hormones related to the menstrual cycle, although the biological basis for this is still not understood (4).

The association of oral contraceptive use and hormone replacement therapy (HRT) with BC risk provides further evidence for the role of ovarian hormones in BC. A systematic review that included 44 BC studies showed that oral contraceptive use increased the risk of BC (5). A large-scale meta-analysis combining case-control data from 58 studies found that HRT use was associated with an increased risk of BC within four years of current use, with increasing risk associated with longer duration of current use (6).

Analyses looking specifically at blood levels of nine sex steroid hormones and BC risk concur with evidence surrounding factors associated with the menstrual cycle, oral contraceptive and HRT use with BC risk. A pooled analysis of nine prospective studies on 663 BC cases and 1765 controls found that increasing concentrations of estrone, androstenedione, dehydroepiandrosterone (DHEA), dehydroepiandrosterone sulfate (DHEAS) and testosterone were associated with increased risk of BC in postmenopausal women (7). Whilst most of these associations were thought to be due to the conversion of androgens (DHEA, DHEAS, testosterone and androstenedione (8,9)) to estradiol, these associations remained even after adjustment for circulating estradiol levels (7,10). Androgen receptors have been shown to increase proliferation when expressed in triple negative breast cancer (TNBC), further providing evidence for a role of androgens in BC risk independent of estradiol (11). A much larger study conducted in UK Biobank among 30,565 pre-menopausal women and 133,294 post-menopausal women found that testosterone and sex hormone-binding globulin (SHBG) increased and decreased BC risk in post-menopausal women, respectively, but did not influence pre-menopausal BC risk (12). Whilst observational evidence has shown associations between sex hormone levels and BC risk, conversely, no association has, as yet, been found between estradiol, testosterone, DHEAS and SHBG and BC survival (13).

Common metabolic pathways may underlie the relationship between sex hormones and BC risk. They are all produced from cholesterol and are synthesised in the gonads, adrenal cortex and the placenta (14). Cholesterol is first transported into the mitochondrion and converted to pregnenolone-the precursor for all sex hormones (Figure 1) (15–17). Whilst approximately half of testosterone originates from the adrenal glands and the ovaries, the remainder is derived from the conversion of proandrogens (DHEA, DHEA-S and androstenedione) in the periphery (18). In post-menopausal women, the primary source of estradiol is from the conversion of androgens (19).

**Figure 1.**
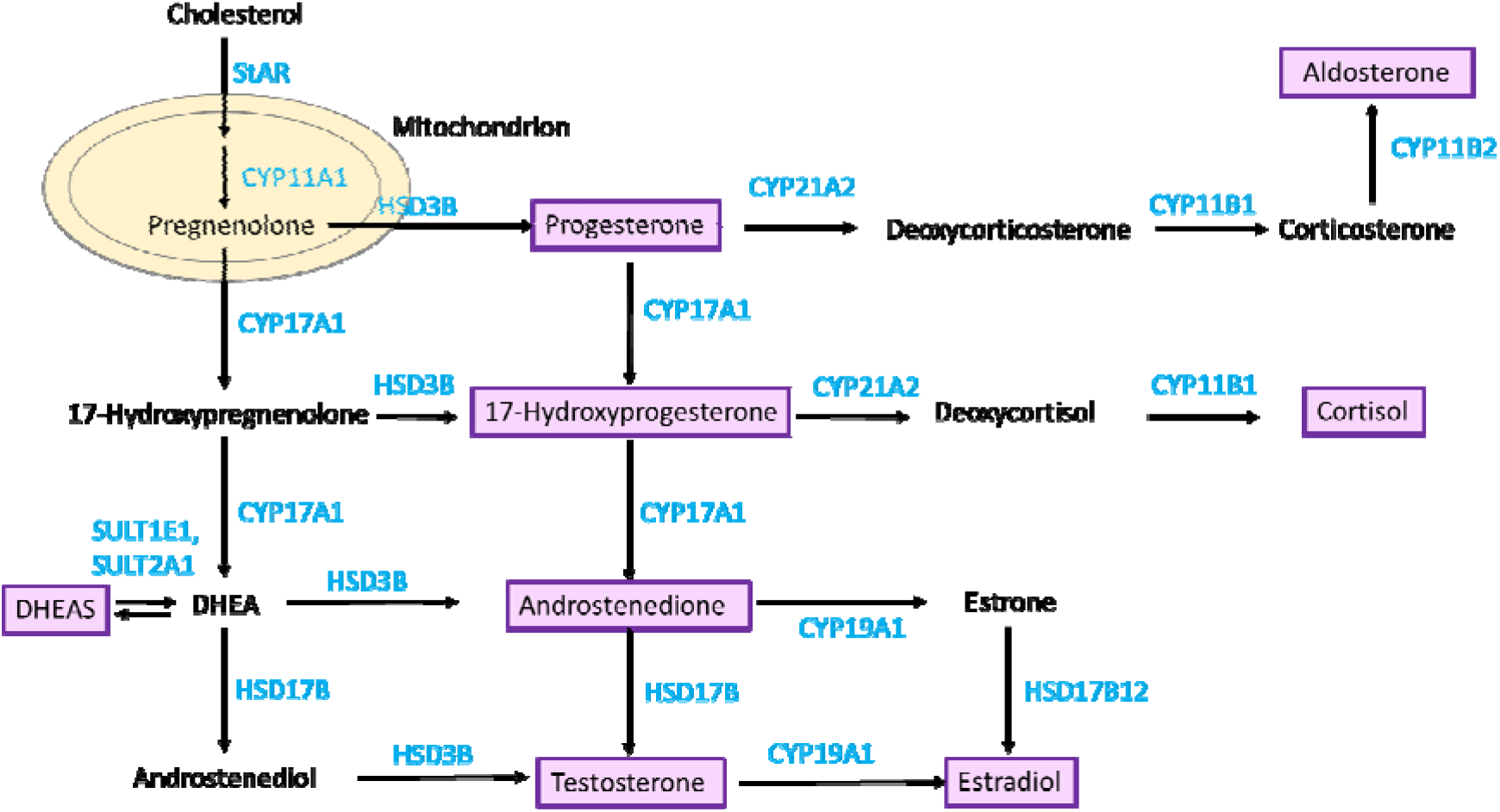
Sex steroid hormone metabolism pathway. Metabolites/hormones are displayed in black text and the enzymes that catalyse the reaction are in blue text. The hormones that are investigated in this analysis are shown in purple boxes. This diagram was adapted from Pott *et al*. (2019) (29).

Much of the current evidence surrounding sex hormones and BC risk comes from observational epidemiological studies. However, these studies are prone to confounding, reverse causation and other biases (20,21). The most reliable method for evaluating the effects of sex hormones on BC risk is through conducting randomized controlled trials (RCTs) but these are time consuming and costly (22), especially in the case of primary prevention trials of cancer. For this reason, other approaches to causal inference such as Mendelian randomization (MR) can be used to provide evidence for or against the role of sex hormones. MR uses genetic variants that predict changes in exposures (e.g. hormone levels) and assesses their effect an outcome (e.g. BC) (21,23,24). MR is analogous to an RCT as genetic variants are randomly allocated at conception, similar to random allocation of intervention at the start of a trial (25,26), and fixed thereafter. This reduces the impact of confounding and reverse causation encountered in observational epidemiology (25).

Previous MR studies have been carried out looking at the effect of testosterone (total and bio-available) and sex hormone binding globulin (SHBG) on overall, ER+ and ER- BC risk (27,28). In this two-sample MR study, we expanded the analysis to include seven other sex steroid hormones as well as investigating the effect of the hormones on subtype specific BC risk (luminal A-like BC, luminal B-like BC, luminal B/HER2-negative-like BC, HER2-enriched-like BC, TNBC and *BRCA1* mutated TNBC) and overall, ER+ and ER- BC survival.

## Materials and Methods

### Two-sample MR

To investigate the effect of sex hormone levels on BC risk and survival, we applied a two-sample MR approach (30). Firstly, genetic instruments to proxy for nine hormones and SHBG were obtained from GWAS summary statistics (sample 1). These were then integrated with BC risk or survival genetic association effect estimates from published GWAS results (sample 2).

### Genetic predictors for sex hormones

Single nucleotide polymorphisms (SNPs) were identified in relation to: total testosterone, bioavailable testosterone, DHEAS, estradiol, cortisol, androstenedione, aldosterone, progesterone, 17-hydroxyprogesterone (17OHP) and SHBG. The threshold for SNP selection was P < 5 × 10 ^-8^. When no SNPs were found at this association, the threshold was relaxed to P < 5 × 10 ^-7^, as was the case for estradiol, aldosterone and androstenedione (Table 1).

**Table 1.** Exposure datasets

| Hormone | Sample N | SNPs N | Year | Instruments N | Units | P-value selection threshold | Variance explained (%) | F statistic | First Author and Reference |
| --- | --- | --- | --- | --- | --- | --- | --- | --- | --- |
| Total testosterone | 230,454 | 16,580,850 | 2020 | 204 | SD | $5 \times 10^{-8}$ | 2.36 | 27.29 | Ruth (27) |
| Bioavailable testosterone | 188,507 | 16,585,744 | 2020 | 131 | SD | $5 \times 10^{-8}$ | 1.77 | 25.86 | Ruth (27) |
| SHBG | 189,473 | 16,585,865 | 2020 | 200 | SD | $5 \times 10^{-8}$ | 3.35 | 32.81 | Ruth (27) |
| DHEAS | 9,722 | 21,770,677 | 2017 | 4 | Log-transformed unit ($\mu\text{mol/L}$) (converted to SD) | $5 \times 10^{-8}$ | NA | NA | Prins (38) |
| Estradiol | 2,607 | 7,705,454 | 2019 | 2 | Log-transformed unit ($\text{pmol/L}$) (converted to SD) | $5 \times 10^{-7}$ | 0.64 | 8.40 | Pott (29) |
| Androstenedione | 711 | 8,799,744 | 2019 | 1 | Log-transformed unit ($\text{nmol/L}$) (converted to SD) | $5 \times 10^{-7}$ | 0.44 | 3.10 | Pott (29) |
| Aldosterone | 685 | 8,806,555 | 2019 | 1 | Log-transformed unit ($\text{pmol/L}$) (converted to SD) | $5 \times 10^{-7}$ | 0.55 | 3.79 | Pott (29) |
| Cortisol | 25,314 | 8,452,426 | 2021 | 1 | SD | $5 \times 10^{-8}$ | 0.11 | 27.07 | Crawford (39) |
| Progesterone | 1,259 | 8,799,744 | 2019 | 3 | Log-transformed unit (nmol/L) (converted to SD) | $5 \times 10^{-8}$ | 0.64 | 2.71 | Pott (29) |
| 17OHP | 711 | 8,799,744 | 2019 | 1 | Log-transformed unit (nmol/L) (converted to SD) | $5 \times 10^{-8}$ | 0.11 | 0.78 | Pott (29) |

SNPs predicting levels of total testosterone, bioavailable testosterone and SHBG were obtained from publicly available summary statistics provided by Ruth *et al*. using UK Biobank data (27), which consists of phenotype and biological samples collected from around 500,000 individuals across the United Kingdom (31). Testosterone and SHBG levels were measured (nmol/L) using a one step competitive analysis and two-step sandwich immunoassay analysis, respectively,on a Beckman Coulter Unicel Dxl 800 in 230,454 and 189,473 participants respectively (27). Bioavailable testosterone (nmol/L; female N: 188,507) was calculated from total testosterone and albumin, which was also measured by BCG analysis on a Beckman Coulter AU5800 (g/L). Genotyping and imputation (using the Haplotype Reference Consortium (HRC) and 1000 Genomes) and quality control (removal of SNPs with MAF ≤ 0.01) filtering resulted in 16,580,850-16,585,865 SNPs for the 3 measures (Table 1) (27). Measures were subjected to inverse normal transformation of rank before being taken forward in a sex-stratified GWAS study (27).

Estradiol has also been measured in UK Biobank using a Beckman Coulter DXI 800 with a detection range between 73-17,621 pmol/L (32). However, since the majority of females (162,691/273,455 (59.49%)(33), mean age:56.35 years (34)) enrolled in the UK Biobank were postmenopausal, levels of estradiol were below the limit of detection for 75% of these women (27,35). Estradiol levels were also measured in the LIFE-Adult and LIFE-Heart cohorts. The LIFE-Adult cohort consists of a random selection of 10,000 participants from Leipzig, Germany. Conversely, 7,000 participants were chosen for the LIFE-Heart study based on having suspected or confirmed coronary artery disease. In contrast to UK Biobank, estradiol measurements in the LIFE-Adult and LIFE-Heart cohort (total N: 2,607) were measured using an electrochemiluminescence immunoassay (ECLIA) with a detection limit of 18.4 pmol/L (36) and liquid chromatography-tandem mass spectrometry (LC-MS/MS) with a lower detection limit of 37 pmol/L, respectively (37). For this reason, we used these two cohorts to obtain genetic instruments to proxy for estradiol levels. The mean age of women in the LIFE-Adult and LIFE-Heart cohorts were 59.4 and 64.8 years, respectively (29). Imputation in this GWAS was performed using 1000 Genomes Phase 3 as the reference panel. Estradiol measurements were log-transformed prior to analysis and so SNP associations represent a log-transformed unit increase (pmol/L) in levels (Table 1) (29).

The hormones androstenedione, aldosterone and 17-OHP were measured in females in LIFE-Heart only (N=711, N=685 and N=711, respectively) (Table 1) using LC-MS/MS. Progesterone was measured in females in both LIFE-Heart and LIFE-Adult using LC-MS/MS (N=1,259). These GWASs were imputed using the 1000 Genomes reference panel with further information in Table 1. Steroid hormone measurements were log-transformed prior to analysis and so SNP associations represent a log-transformed unit increase (nmol/L or pmol/L) in hormone levels (Table 1) (29).

We obtained summary statistics for DHEAS associations from Prins *et al*. which included DHEAS measures for 9,722 participants (4,308 males and 5,414 females) obtained from the United Kingdom Household Longitudinal Study - a longitudinal survey across the UK (England, Wales, Scotland and Northern Ireland) consisting of 40,000 households (38). DHEAS was measured (µmol/L) in serum samples using a competitive immunoassay on the Roche E module analyser and measurements were log transformed and adjusted for age and sex, thereby SNPs represented a log-transformed unit (µmol/L) increase in DHEAS levels (38). Imputation was performed using the UK10K project and 1000 Genomes phase 3 panels in this GWAS (Table 1) (38).

We obtained summary statistics for cortisol from the CORtisol NETwork (CORNET) consortium that meta-analysed GWAS statistics from 17 population-based cohorts of a European background including 25,314 individuals (36.27% men and 63.73% women) (39).Cortisol was measured using immunoassays in blood samples for all studies except TwinsUK which used liquid chromatography-mass spectrometry. The studies performed linear regressions on z-scores of log-transformed morning plasma cortisol and were also adjusted for sex, age and genetic ancestry. Imputation was performed using the Haplotype Reference Consortium and 1000 Genomes Phase 3 panels in this GWAS, with SNPs representing an SD increase in cortisol levels (Table 1) (39).

For estradiol, DHEAS, progesterone, 17-OHP, aldosterone and androstenedione, log-transformed units were converted to a standard deviation scale so that results would represent a SD increase in hormone levels. For hormones with reported median and interquartile (IQR) ranges, these were transformed to the log scale and the SD was calculated using the method presented by Wan *et al*. (40). DHEAS study characteristics reported mean, maximum and minimum values, which were transformed to the log scale and the SD was calculated using the formula: (maximum – minimum)/4. When the hormone was measured in more than one study (estradiol, DHEAS and progesterone), a combined SD was calculated using the formula from the Cochrane Handbook (Section 6.5.2.10) (41).

To adjust for multiple testing, we applied a genome-wide significance threshold for SNP associations with metabolites (P value ≤ 5×10 ^-8^). When no SNPs were available at this cut-off, we relaxed the threshold to a P value ≤ 5×10^−7^ (Table 1). We also chose to include independent SNPs to avoid multi-collinearity and therefore carried out linkage disequilibrium (LD) clumping at an R^2^<0.001 so that only the SNP most strongly associated with the hormone within a 10,000kb window was taken forward in the analysis.

We calculated the variance explained as well as the F statistic to assess whether the identified SNPs may be weak instruments. When weak instruments are used in a two-sample MR analysis, the estimate obtained tends to be biased towards the null (42). The F statistic helps to determine the strength of the bias, with lower F statistics indicating greater bias towards the null (43). Power calculations were conducted using the mRnd online calculator to identify the effect size (OR) in both directions that could be detected based on the variance explained by the instruments and the sample sizes available (44). Due to the absence of effect allele frequencies for the DHEAS GWAS, the variance explained, F statistic and power calculations could not be calculated for this hormone.

### Genetic associations for breast cancer

#### Breast cancer risk

Genetic association summary statistics for BC risk were obtained from the Breast Cancer Association Consortium (BCAC) (consisting of 68 studies combined together) as well as the Discovery, Biology and Risk of Inherited Variants in Breast Cancer Consortium (DRIVE) (45). This study includes 122,977 BC cases and 105,974 controls and when stratified based on estrogen receptor (ER) expression, there were 69,501 ER+ BC cases and 21,468 ER- BC cases (Table 2). Genotyping was carried out using both the iCOGs array or the OncoArray with imputation (using the version 3 release of the 1000 Genomes Project dataset as a reference panel) to obtain data on 10,680,257 SNPs and results were combined using a fixed-effect meta-analysis (45).

**Table 2.**
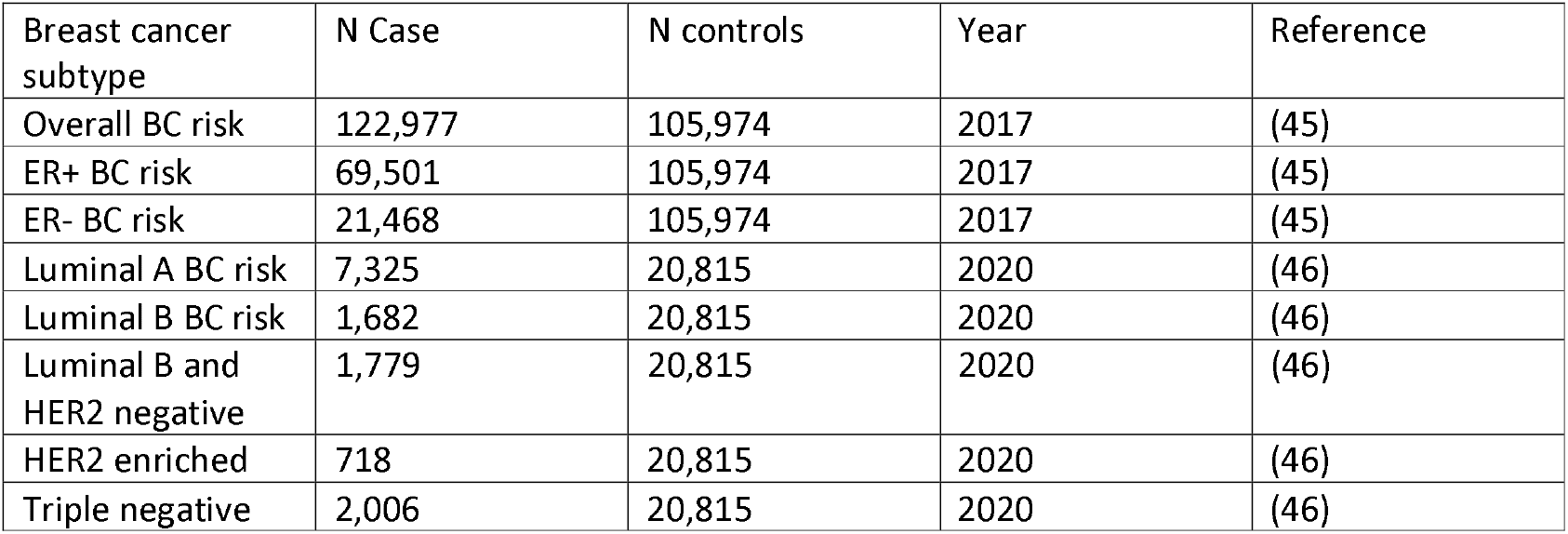
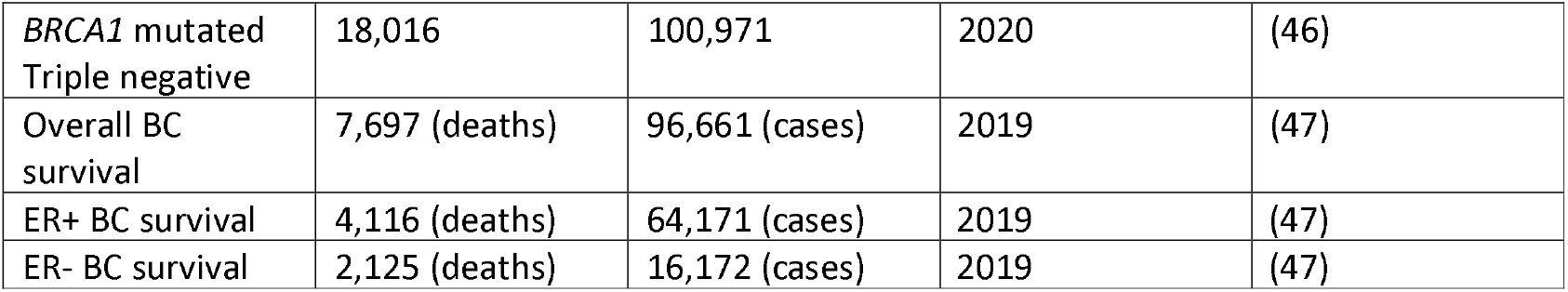
Outcome datasets

#### Breast cancer risk subtypes

To further identify subtype specific effects of hormones on BC risk, we also tested their association with 6 subtypes of BC. Data from 118,474 cases and 96,201 controls were previously analysed from 82 studies from BCAC to obtain summary statistics associations for 5 subtypes of BC. These subtypes are defined by expression of ER, PR, HER2 and cancer grade: luminal A-like (ER+, and/or PR+, HER2- and grades 1/2 ; 7,325 cases and 20,815 controls), luminal B-like (ER+ and/or PR+, HER2+; 1,682 cases and 20,815 controls), luminal B/HER2-negative-like (ER+ and/or PR+, HER2-, grade 3; 1,779 cases and 20,815 controls), HER2-enriched-like (ER-, PR-, HER2+; 718 cases and 20,815 controls) and triple-negative breast cancer (TNBC) (ER-, PR-, HER2-; 2,006 and 20,815 controls) (46). Furthermore, summary data from the Consortium of Investigators of Modifiers of BRCA1/2 (CIMBA) comprising 9,414 cases with *BRCA1* mutation and 9494 controls with *BRCA1* mutation was also used in this analysis. Since the majority of *BRCA1*-mutated cancers were also triple-negative, we used summary statistics that meta-analysed associations of *BRCA1*-mutated cancers with TNBC (18,016 cases and 100,971 controls) (Table 2) (46).

#### Breast cancer survival

To investigate the role of hormones in BC survival, we used summary statistics that consisted of 12 studies including 96,661 women with BC and 7,697 BC-specific deaths, of which details for each study have been described previously (47). Analyses were also stratified based on tumour ER expression, with 64,171 ER+ BC women and 4,116 ER+ BC-specific deaths as well as 16,172 ER-negative BC patients and 2,125 ER- BC-specific deaths (Table 2) (47). Cox proportional hazards regression were used to calculate the hazard ratios (HR) of genotype association with BC-specific mortality (47).

### Statistical analysis

Analyses were carried out in R version 3.3.1 using the “Two-Sample MR” package (48), which allows data formatting, harmonisation and application of MR methods in a semi-automated manner. This package automatically assigns the allele with a positive effect on the exposure as the effect allele, so that the effect allele predicts an increase in hormone levels. The SNPs used to proxy for the exposure are also extracted from the BC outcome datasets. Exposure and outcome summary statistics are then subject to allele harmonization to ensure that the effect allele in the exposure dataset (hormone-increasing) is the same effect allele in the outcome dataset, with effect allele frequencies used to assist harmonizing palindromic SNPs.

In the presence of one SNP to proxy for hormone levels, Wald ratios (SNP-outcome estimate/SNP-exposure estimate) were used to calculate the change in log OR (risk analysis) or log HR (survival analysis) per SD increase in hormone levels. When more than one SNP was present, an inverse variance weighted (IVW) (fixed effects) method was applied, which is an average of the Wald ratios where the weight of the SNP contribution to the overall estimate is the inverse of the SNP effect on the outcome (49,50). We performed the analysis for the risk of overall BC, ER+ BC, ER- BC, luminal A-like BC, luminal B-like BC, luminal B/HER2-negative-like BC, HER2-enriched-like BC, TNBC and *BRCA1* mutated TNBC. We also performed the analysis for survival of overall BC, ER+ BC and ER- BC.

The IVW method is prone to bias if one of the genetic instruments is invalid due to its association with another trait through an independent pathway (horizontal pleiotropy) (51). For this reason, we also applied alternative MR methods that produce unbiased estimators even in the presence of some invalid genetic instruments. When more than 2 SNPs were present, we calculated a weighted median, weighted mode and an MR Egger estimate (48,52–54). The weighted median approach allows a consistent estimate even if 50% of the information contributing to the overall estimate comes from invalid genetic instruments (53). The weighted mode estimate may also be used even when the majority of the SNPs are invalid instruments so long as the SNPs that form a cluster of homogenous results are valid (49,52). Finally, we adopted an MR Egger analysis to evaluate evidence for the presence of horizontal pleiotropy. This method is not constrained to pass through an effect size of 0, therefore the y intercept gives an indication of the presence of directional pleiotropy (51,54). We used an MR Egger intercept with a P value below 0.05 to indicate the presence of directional pleiotropy that may be influencing the MR results. For hormones proxied by weak instruments, we conducted an MR robust adjusted profile score (MR RAPS), a method that provides robust inference when weak instruments are present (44).

Linkage disequilibrium score regression (LDSC) was used to assess the genetic correlation between total testosterone and bioavailable testosterone with estradiol using the settings advised in the software package LDSC (v1.0.1) (55). We tested these hormones due to the direct conversion of testosterone to estradiol and therefore to identify whether SNP associations are shared between the 2 traits. Firstly, quality control was performed on the summary statistics to exclude variants with missing data, non-biallelic, strand-ambiguous alleles which could not be matched in the European ancestry 1000 Genomes reference panel, variants with imputation scores below 0.90 and rare variants with minor allele frequencies below 0.01. A score was calculated to reflect whether GWAS test statistics for variants correlates with nearby variants that are in high LD. A z statistic was generated for each variant in trait 1 and this was multiplied with the z statistic from trait 2. The product was regressed against the LD scores and the resultant coefficient/slope was the genetic correlation statistic.

## Results

### Two-sample MR analysis of sex hormones and breast cancer risk

To investigate the effect of sex hormones on BC risk, we conducted an MR analysis of nine hormones and SHBG on overall, ER+ and ER- BC risk.

After clumping at R^2^<0.001, we identified 204 and 131 SNPs to proxy for a SD increase in total testosterone and bioavailable testosterone at P ≤ 5×10^−8^, respectively (Supplementary Table 1). These SNPs explain 2.36% and 1.77% of the variance in the hormone levels and have an F statistic of 27.29 and 25.87, respectively (Supplementary Table 2). Using an IVW approach, we found that an SD increase in total testosterone increased the risk of overall BC and ER+ BC (OR: 1.14, 95% CI: 1.09-1.21 and OR: 1.19, 95% CI: 1.12-1.27, respectively) but had no effect on ER- BC (OR: 0.99, 95% CI: 0.93-1.06) (Figure 2). We also found positive associations between total testosterone and overall BC risk using the MR Egger, weighted median and weighted mode methods (OR:1.22, 95% CI: 1.10-1.34, OR:1.13, 95% CI: 1.06-1.20 and OR:1.17, 95% CI: 1.08-1.26, respectively) as well as a positive association with ER+ BC risk (MR Egger OR: 1.29, 95% CI: 1.16-1.45, weighted median OR: 1.17, 95% CI: 1.09-1.27 and weighted mode OR: 1.23, 95% CI: 1.13-1.35) (Supplementary Table 3). Furthermore, we conducted an MR Egger intercept test but found no evidence of directional pleiotropy for overall BC risk and ER+ BC risk (Supplementary Table 4).

**Figure 2.**
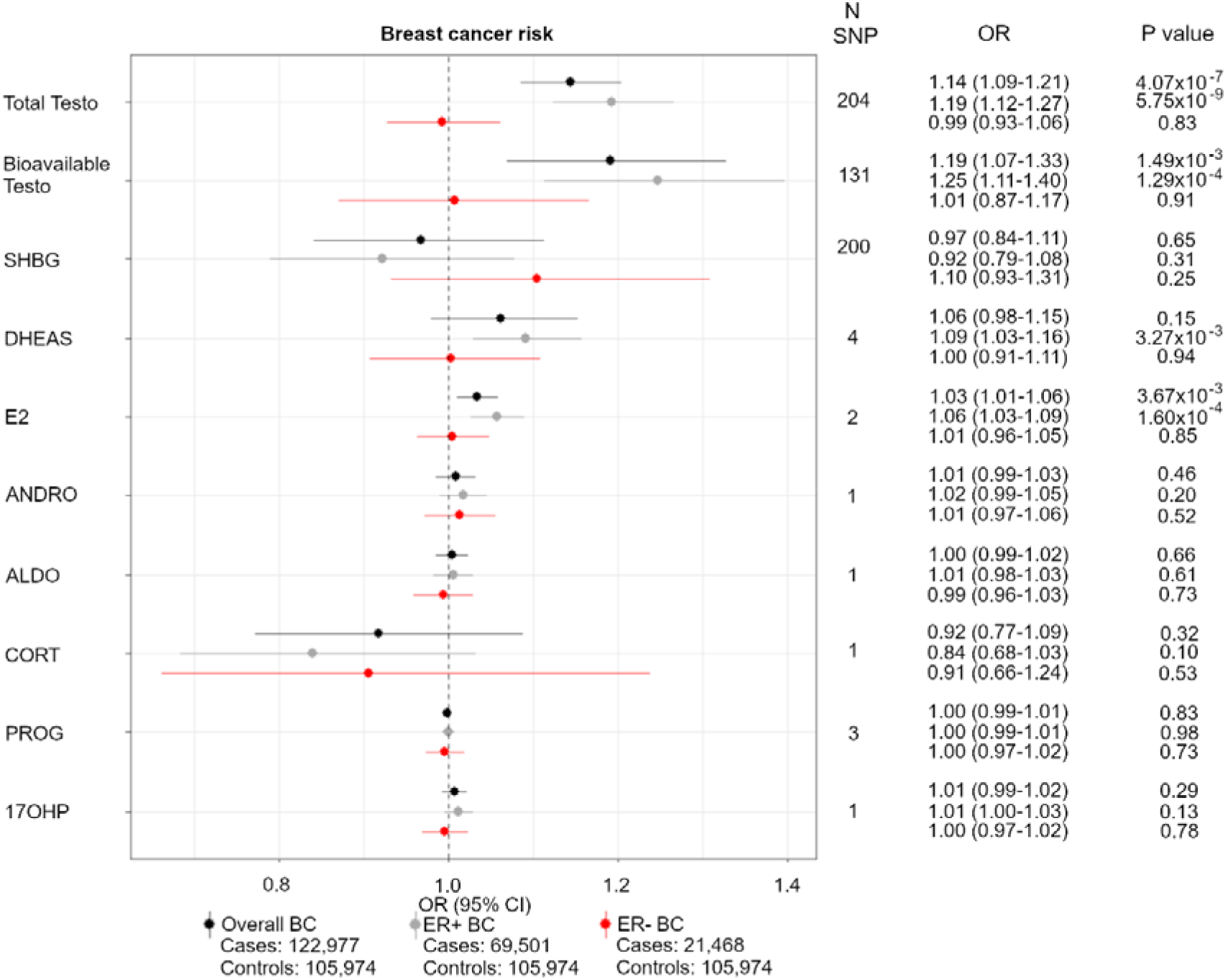
Forest plot showing the MR associations between sex hormones and overall, ER+ and ER- BC risk. IVW analysis was carried out to assess the association between an SD increase in total testosterone, bioavailable testosterone, SHBG, DHEAS, estradiol, androstenedione, aldosterone, cortisol, progesterone and 17-OHP on risk of incidence of overall BC (black), ER+ BC (grey) and ER- BC (red). Abbreviations: OR, odds ratio, Total Testo, total testosterone; Bioavailable Testo, bioavailable testosterone; SHBG, sex hormone-binding globulin; DHEAS, dehydroepiandrosterone sulphate; E2, estradiol; ANDRO, androstenedione; ALDO, aldosterone; CORT, cortisol; PROG, progesterone; 17OHP, 17-hydroxyprogesterone.

We also found that a SD increase in bioavailable testosterone increased the risk of overall and ER+ BC (OR: 1.19, 95% CI: 1.07-1.33 and OR: 1.25, 95% CI: 1.11-1.40, respectively). Further positive associations were found using the weighted median approach (overall BC risk: 1.13, 95% CI: 1.02-1.24 and ER+ BC OR: 1.23, 95% CI: 1.10-1.38) and suggestive positive associations from the MR Egger and weighted mode approaches (Supplementary Table 3).

We found 2 SNPs at P≤5×10^−7^ to proxy for an SD increase in estradiol, which explained 0.64% of the variance with an F statistic of 8.40. Although the F statistic is low, one of the SNPs (rs2414098) is an intronic variant in the gene *CYP19A1* (56) which encodes the enzyme involved in conversion of testosterone to estradiol (29). For this reason, we continued with the MR analysis and found an increased risk of both overall BC risk and ER+ BC (OR: 1.03, 95% CI: 1.01-1.06 and OR: 1.06, 95% CI: 1.03-1.09, respectively) but no effect on ER- BC risk (OR: 1.01, 95% CI: 0.96-1.05).

Due to the possibility of weak instrument bias, we also used the MR-RAPS method and found a positive association between a SD increase in estradiol with overall and ER+ BC (OR: 1.04, 95% CI: 1.01-1.06 and OR: 1.06, 95% CI: 1.02-1.09, respectively). Since only 2 SNPs were used as proxies for estradiol, no other MR methods were used to test this association. We also calculated the genetic correlation between total testosterone and bioavailable testosterone with estradiol, but found no strong evidence of correlation (total testosterone r_*g:*_ 0.25 (95% CI: -0.05-0.54) and bioavailable testosterone r_*g:*_ 0.09, (95% CI: -0.13-0.31)) (Supplementary Table 5), indicating that the effect of estradiol on BC risk may be independent of testosterone levels.

Similar to total testosterone, bioavailable testosterone and estradiol, we found a positive association between a SD increase in DHEAS (proxied by 4 SNPs) and ER+ BC risk (OR: 1.09, 95% CI:1.03-1.16), with positive associations also observed using the weighted median, weighted mode and MR-Egger approaches (OR: 1.08, 95% CI: 1.04-1.13, OR: 1.08, 95% CI: 1.03-1.13, OR: 1.07, 95% CI: 0.97-1.18, respectively) (Supplementary Table 3).

We found little evidence of association between SHBG or cortisol levels and overall, ER+ and ER- BC risk. With regards to androstenedione, aldosterone, progesterone and 17-OHP, we also found little evidence of associations with overall, ER+ and ER- BC risk. However, we acknowledge that the genetic instruments used to proxy these hormones may be weak as demonstrated by low F statistics (0.78-3.79) (58) (Table 1).For this reason, we also carried out a weak instrument robust method – MR-RAPS, but still found little evidence of an association between the four hormones and overall, ER+ and ER- BC risk (Supplementary Table 3).

### Two-sample MR analysis of sex hormones and breast cancer subtype risk

To investigate the effect of sex hormones on the risk of specific BC subtypes, we conducted an MR analysis of the nine hormones and SHBG on luminal A-like BC, luminal B-like BC, luminal B/HER2-negative-like BC, HER2-enriched-like BC, TNBC and *BRCA1*-mutated TNBC. Details on the SNPs used as instruments in this analysis are found in the Supplementary Tables 6 and 7.

We found that a SD increase in total testosterone levels increased the risk of luminal A-like BC, luminal B-like BC and luminal B/HER2-negative-like BC (OR: 1.21, 95% CI: 1.13-1.28, OR: 1.14, 95% CI: 1.02-1.26 and OR: 1.21, 95% CI: 1.11-1.31, respectively) (Figure 3). Similar directions of association were found using the weighted median, weighted mode and MR-Egger approaches (Supplementary Table 3) with the MR-Egger intercept showing no evidence of directional pleiotropy (Supplementary Table 4). Conversely, a SD increase in total testosterone was associated with a decreased risk of *BRCA1*-mutated TNBC (OR: 0.91, 95% CI: 0.84-0.99). We found positive associations between a SD increase in bioavailable testosterone and luminal A-like BC and luminal B/HER2-negative-like BC risks (OR: 1.29, 95% CI: 1.15-1.43 and OR: 1.22, 95% CI: 1.07-1.40) with consistent directions of association found using the weighted median, weighted mode and MR-Egger approaches for the association with luminal A-like BC. However, only the MR-Egger approach showed a positive association for luminal B/HER2-negative-like BC risk (Supplementary Table 3). In contrast, the MR Egger intercept showed little evidence of directional pleiotropy for any of these associations (Supplementary Table 4).

**Figure 3.**
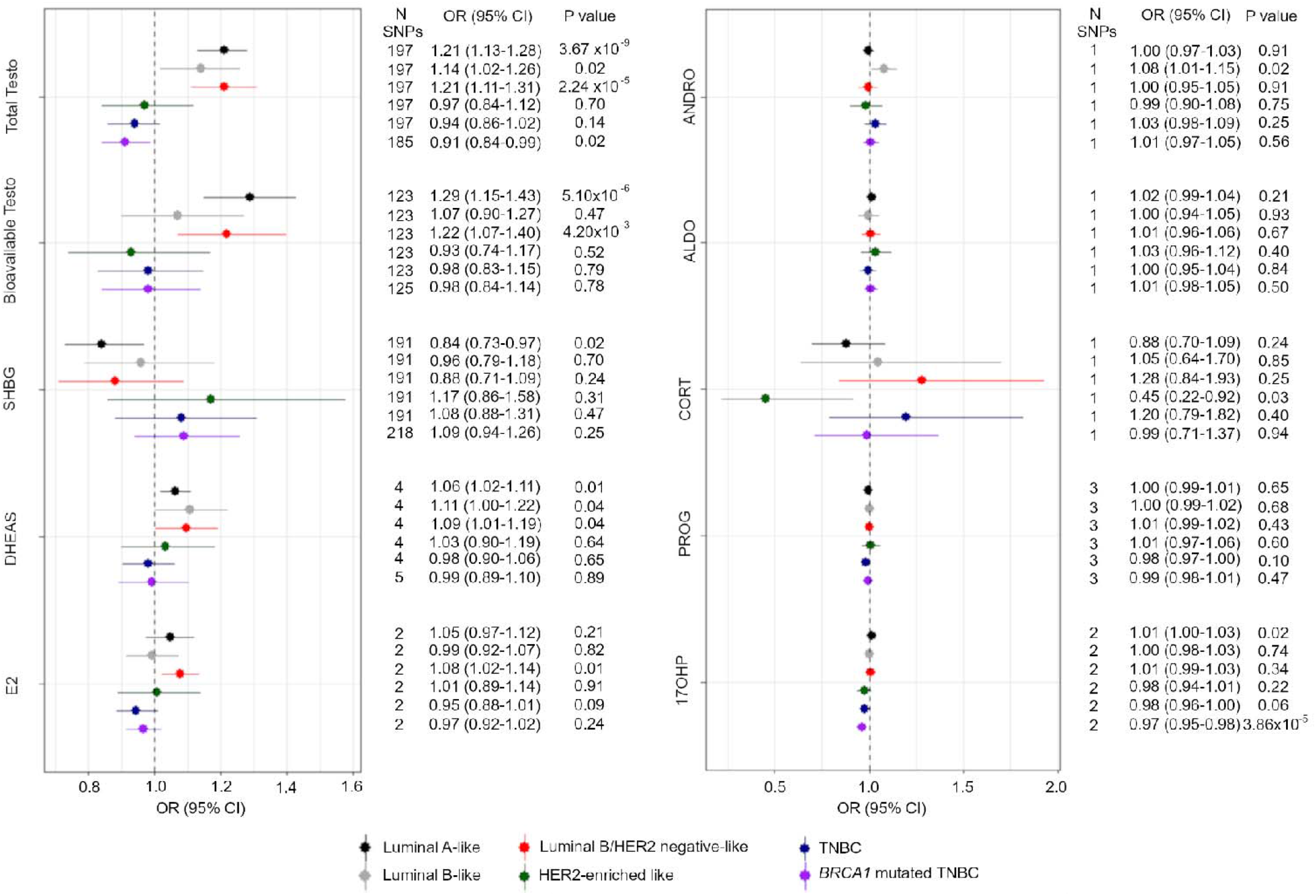
Forest plot showing the MR associations between sex hormones and risk of six BC subtypes. IVW analysis was carried out to assess the association between an SD increase in total testosterone, bioavailable testosterone, SHBG, DHEAS, estradiol, androstenedione, aldosterone, cortisol, progesterone and 17-OHP on risk of Luminal A BC (black), Luminal B (grey), Luminal B and HER2 negative BC (red), HER2 enriched BC (green), Triple negative BC (blue) and BRCA1 mutated triple negative BC (purple). Abbreviations: OR, odds ratio; Total Testo, total testosterone; Bioavailable Testo, bioavailable testosterone; SHBG, sex hormone-binding globulin; SHBG adjusted, SHBG adjusted for BMI; DHEAS, dehydroepiandrosterone sulphate; E2, estradiol; ANDRO, androstenedione; ALDO, aldosterone; CORT, cortisol; PROG, progesterone; 17OHP, 17-hydroxyprogesterone; TNBC, triple negative BC.

We found an inverse association between an SD increase in levels of SHBG and luminal A-like BC risk (OR: 0.84, 95% CI: 0.73-0.97), however, the weighted median, weighted mode and MR Egger approaches showed little evidence of association (Supplementary Table 3).

Similar to total testosterone and bioavailable testosterone, we found that a SD increase in estradiol increased the risk of luminal B/HER2-negative-like BC (OR: 1.08, 95% CI: 1.02-1.14) and a possible inverse association with the more aggressive subtype of cancer (TNBC IVW OR: 0.95, 95% CI: 0.88-1.01) (Figure 3). We also found similar associations using the MR-RAPS method (OR:1.08, 95% CI: 1.01-1.15 and OR: 0.94, 95% CI: 0.89-1.00). Due to a limited number of SNPs, further MR methods could not be applied.

We also found positive associations between a SD increase in DHEAS and luminal A-like BC, luminal B-like BC and luminal B/HER2-negative-like BC (OR: 1.06, 95% CI: 1.02-1.11, OR: 1.11, 95% CI: 1.00-1.22 and OR: 1.09, 95% CI: 1.01-1.19, respectively) and found concurring results with the weighted median approach for all subtypes and suggestive positive associations for both the weighted mode and MR-Egger approaches (Supplementary Table 3). We saw little evidence of directional pleiotropy from the MR-Egger intercept (Supplementary Table 4).

With regards to cortisol, we found an inverse association between a SD increase in the hormone and HER2-enriched-like BC (OR: 0.45, 95% CI: 0.22-0.92). The F statistics and variance explained for the remaining four hormones (androstenedione, aldosterone, progesterone and 17-OHP) were suggestive of weak instruments and power calculations showed that we were underpowered to detect an effect for some of the BC subtypes (Supplementary Table 8). We found a positive association between androstenedione and luminal B-like BC using the IVW method (OR: 1.08, 95% CI: 1.01-1.15) but this effect was no longer observed using the weak instrument-robust MR-RAPS method (OR: 1.02, 95% CI: 0.99-1.05). The MR-RAPS method did suggest a possible association of androstenedione with TNBC (OR: 1.08, 95% CI: 1.00-1.17). We found little evidence of associations between aldosterone and progesterone with risk of the six BC subtypes using both the IVW and MR-RAPS method. Finally, we observed a positive association between a SD increase in 17-OHP and luminal A-like BC (OR: 1.01, 95% CI: 1.00-1.03) and an inverse association with *BRCA1*-mutated TNBC (OR: 0.97, 95% CI: 0.95-0.98) using the IVW method. Both of these associations were also observed when using the MR-RAPS method (luminal A-like BC OR: 1.01, 95% CI: 1.00-1.03 and *BRCA1*-mutated TNBC OR: 0.97, 95% CI: 0.95-0.99).

### Two-sample MR of sex hormones and breast cancer survival

We further conducted an MR analysis to investigate the effect of sex hormones on BC survival (details on SNPs used as instruments can be found in Supplementary Table 9). In contrast to the association between total testosterone or bioavailable testosterone and BC risk, we found little evidence of an association between both hormones and BC survival, regardless of tumour ER status (Figure 4). We also found little evidence of an association between SHBG, DHEAS, estradiol, androstenedione or aldosterone and BC survival. Of all the hormones, only a SD increase in cortisol was associated with a worse outcome of ER- BC survival (HR: 2.35, 95% CI: 1.00-5.49).

**Figure 4.**
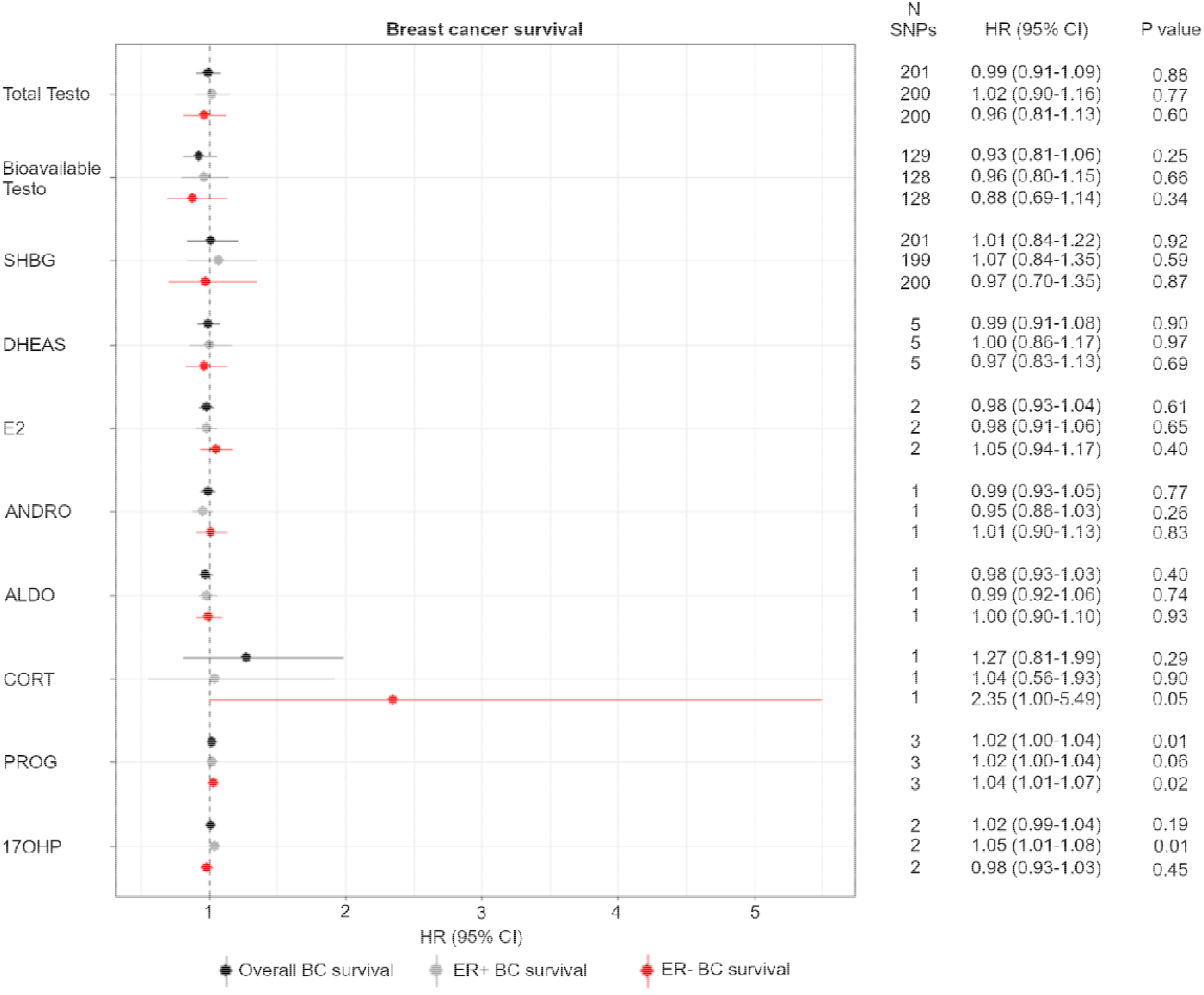
Forest plot showing the MR associations between sex hormones and overall, ER+ and ER- BC survival. IVW analysis was carried out to assess the association between an SD increase in testosterone, DHEAS, estradiol, androstenedione, aldosterone, cortisol, progesterone and 17-OHP on overall BC survival (black), ER+ BC (grey) and ER- BC (red). Abbreviations: HR, hazard ratio; TESTO, testosterone; DHEAS, dehydroepiandrosterone sulphate; E2, estradiol; ANDRO, androstenedione; ALDO, aldosterone; CORT, cortisol; PROG, progesterone; 17OHP, 17-hydroxyprogesterone; BC, breast cancer.

With regards to progesterone, we observed a weak positive association between a SD increase in the hormone and overall BC, ER+ and ER- BC survival (HR: 1.02, 95% CI: 1.00-1.04, HR: 1.02, 95% CI:1.00-1.04 and OR: 1.04, 95% CI: 1.01-1.07, respectively). Due to the possibility of weak instrument bias, we also assessed the association using MR-RAPS and found a consistent positive association between progesterone with overall and ER- BC survival (HR: 1.02, 95% CI: 1.00-1.04 and HR: 1.04, 95% CI: 1.00-1.07, respectively). Furthermore, we observed a positive association between a SD increase in 17-OHP and ER+ BC survival (HR: 1.05, 95% CI: 1.01-1.08) which was also observed using the MR-RAPS method (HR: 1.05, 95% CI: 1.01-1.08).

## Discussion

In this study, we aimed to assess whether nine sex steroid hormones and SHBG affect BC risk and survival using an MR framework. Overall, we found that an increase in testosterone, bioavailable testosterone, estradiol and DHEAS were associated with overall BC and/or ER+ BC. We also found little evidence to suggest an association between the hormones and BC survival, except for a possible worse outcome in terms of ER- BC survival for elevated cortisol levels.

The association of total testosterone and bioavailable testosterone with overall BC as well as ER+ BC risk has been reported before using MR methods (27). However, our study also investigated subtype specific associations using MR in ER+ tumours luminal-A like, luminal-B like and luminal B/HER2-negative-like BC. Associations with ER+ tumours involving testosterone may be explained by two possible mechanisms - the first involves its conversion to estradiol (19) which binds to the ER and induces transcription of growth-positive genes and reduces expression of negative regulators of cell growth, therefore increasing breast cancer cell proliferation (59). The second possible explanation for the association may be due to ER expression acting as a proxy for androgen receptor (AR) expression of which AR expression is positively correlated with ER expression in tumours (60,61). This is further supported by the finding that only 20-30% of ER- BCs express AR (62). The literature suggests that resistance to ER therapies may be due to tumour adaptation towards androgen dependence and AR signalling instead and it has been suggested that patients with ER+/AR+ tumours would most likely benefit from combination therapies targeting both receptors (63). In order to try and untangle the mechanism through which testosterone acts in breast cancer, genetic association studies on tumour subtypes stratified based on AR expression and ER expression are required.

Our study demonstrated a relationship between estradiol and BC risk in an MR framework. An SD increase in estradiol increased the risk of overall BC and ER+ BC as well as the ER+ BC subtype luminal B/HER2-negative-like. We used summary statistics from Pott *et al*. to identify suitable genetic instruments for estradiol (29). Measures of E2 were carried out using LC/MS and ECLIA for the LIFE-Adult and LIFE-Heart cohort studies, respectively (29). These two methods are more sensitive than the Beckman Coulter DXI 800 used in UK Biobank, whereby E2 levels could not be detected in the majority of women (27). For this reason, despite the smaller sample size, we used summary statistics from the E2 GWAS by Pott *et al*. instead (29). Despite the more sensitive methods for measuring E2, the average levels of the hormones detected for LIFE-Adult and LIFE-Heart were 18.4 pmol/L and 11.1 pmol/L, respectively(29), compared to ≥200 pmol/L in premenopausal women (29). This may be because the average age of women in these studies was 59.4 and 64.8 years, respectively, which indicates that a large percentage of these cohorts may have been postmenopausal. Postmenopausal women no longer produce estradiol from the ovaries and so production of this hormone is through the conversion of androgens to estradiol which occurs at the tissue of interest (19). Therefore, estradiol production in postmenopausal women is localised and may have resulted in lower detection of estradiol in the blood.

These low levels of estradiol may explain why we only had 2 instruments to proxy for the hormone and why they only explained 0.64% of the variance, with a low F statistic indicating these were weak instruments (8.40). However, one SNP (rs2414098) near *CYP19A1* shows evidence for a key biological role in affecting hormone levels, with suggestive evidence that it is associated with an increase in *CYP19A1* expression in breast tissue and would theoretically result in increased conversion of testosterone to estradiol. This plausible biological role of the SNP supports the results, despite the F statistic suggesting weak instruments. We also find no strong evidence of genetic correlation with total testosterone and bioavailable testosterone, indicating that the estradiol effect on BC risk may be independent of testosterone.

Similarly, an SD increase in DHEAS was associated with an increased risk of ER+ BC, and subtype analysis showed positive associations with the 3 ER+ BC subtypes: luminal A-like, luminal B-like and luminal B/HER2-negative-like BC. These results support positive associations found in observational studies between DHEAS levels and BC risk in postmenopausal women (7,64).

While observational studies have shown generally consistent results with regards to sex hormones and postmenopausal BC risk, few studies have looked at the association with premenopausal BC. ER+ BC is generally found in older and postmenopausal women and ER- BC is generally found in younger premenopausal women (65). Analysis of seven prospective studies found that doubling concentrations of estradiol, androstenedione, DHEAS and testosterone all increased the risk of premenopausal BC (66). Unlike postmenopausal women, premenopausal women produce estradiol in the ovaries which then circulates in the blood (15). The difficulty in trying to understand the relationship between estradiol and BC in premenopausal women is due to the much smaller sample sizes of cases in prospective cohorts as well as difficulty in accounting for the phase of the menstrual cycle which impacts on measures of estradiol (15). For this reason, the association between estradiol and premenopausal BC is still unclear (15). It is important to note that the sex steroid hormone GWASs used in this study have mostly been conducted in older aged women of which a large percentage are post-menopausal. Since ER- BC tends to occur more commonly in pre-menopausal women, instruments robustly predicting hormonal levels in pre-menopausal women need to be identified and used instead.

With regards to BC survival, we found an association between cortisol and a worse outcome of ER- BC survival. Cortisol levels tend to be highest before awakening and then decrease throughout the day. Sephton *et al*. have shown that changes to this pattern have been associated with worse BC survival (67). A lack of association between sex hormones and BC mortality has also been shown in results from two large prospective cohorts of US women with 2,073 breast cancer cases. They found a suggestive association between estradiol and overall BC mortality (HR: 1.43, 95% CI: 1.03-1.97) but no association was observed for testosterone, DHEAS and SHBG (13).

A limitation of conducting an MR analysis using survival GWAS associations is the presence of selection bias. Individuals included in the survival GWAS analysis are all cases and therefore any factors that cause disease incidence will be highly prevalent in this population but would not be so in the general population (68). For example, variants showing an association between testosterone and BC risk may not be associated with confounders in the general population, but may be associated with confounders in a population of BC cases only. By choosing to only include BC cases in the analysis (which is a collider), this means that common causes of BC risk become conditionally associated with each other (69). Therefore, genetic variants that would be distributed randomly in the general population will not be so random in this subpopulation of BC cases which may lead to collider bias (69). Methods are emerging to adjust for this bias and would be useful to conduct before robust conclusions about sex hormones and BC survival can be made (70).

Whilst our study showed associations between testosterone, estradiol, DHEAS and cortisol with various BC subtypes’ risk and survival in an MR framework, it is not without limitations. Firstly, the sample sizes of the GWAS from which some of our exposure instruments were derived were relatively small, and therefore the instruments used were weak, especially in the case of androstenedione, aldosterone, progesterone and 17-OHP. In the case of a two-sample MR setting, using weak instruments will bias the causal estimate towards the null (68) and may explain some of the null associations observed. The lack of genome-wide significant SNPs for androstenedione and aldosterone may have been due to the small sample sizes of the GWASs for these hormone measurements (N=712 and N=686, respectively). In addition, participants in the LIFE-Heart study were selected based on suspected or confirmed coronary artery disease, indicating possible selection bias. Furthermore, we derived genetic instruments for DHEAS and cortisol from mixed populations, due to much larger sample sizes than female-specific GWASs. This means that larger GWASs specifically in females need to be performed to identify stronger genetic instruments for these hormones before definitive conclusions on null associations can be made.

Overall, our results suggest that increasing levels of testosterone, bioavailable testosterone, estradiol and DHEAS may increase the risk of overall BC and/or ER+ (postmenopausal) BC risk, consistent with results from observational studies. For the remaining hormones, we found some suggestive associations but also acknowledge the possibility of weak instrument bias and the need for better genetic instruments. Our study provides new insights into the role of sex steroid hormones in BC risk using MR and may inform the eventual development of interventions aimed at BC prevention.

## Data Availability

All data produced in the present work are contained in the manuscript

